# “It makes you realise your own mortality.” A qualitative study on mental health of older adults in the UK during COVID-19

**DOI:** 10.1101/2020.12.15.20248238

**Authors:** Alison McKinlay, Daisy Fancourt, Alexandra Burton

## Abstract

**Background:** Older adults have been disproportionately affected by COVID-19, with high fatalities and health complications reported. Adults over the age of 70 in the UK were advised to self-isolate for 3 months early during the pandemic and it is unclear which factors influenced their experiences during this time.

**Objective:** The aim of this qualitative study was to explore factors that threatened and protected the wellbeing of older adults living in the UK during the COVID-19 pandemic.

**Methods:** We undertook semi-structured interviews with 20 adults aged over 70. Purposive sampling methods were used to increase diversity within the group. Transcripts were analysed using thematic analysis.

**Results:** Participants were aged 72-93, 9 women and 11 men, 80% were White British, 40% lived alone. We identified 2 superordinate themes, including (1) Threats to wellbeing: mortality concerns, grief and loss of normal life, restricted health service access, COVID-19 concerns, and restricted access to activities that protect wellbeing. (2) Factors protective of wellbeing: slower pace of life, maintaining routine, socialising, and use of past coping skills. Many participants drew on their resilience and life experience to self-manage fear and uncertainty associated with the pandemic, using their time during lockdown to reflect or organise end-of-life affairs.

**Conclusions:** This study provides evidence that while older adults experienced challenges, many were resilient against COVID-19 restrictions despite early concerns of mental health consequences. Our findings highlight the importance of maintaining access to essentials to promote feelings of normality and social support to help reduce uncertainty in times of pandemics.

## 1. Introduction

Existing public health concerns were exacerbated when severe acute respiratory syndrome coronavirus 2 (SARS-CoV-2) was declared a pandemic by the World Health Organisation (WHO) on March 11^th^, 2020 [1]. Older adults were identified as especially vulnerable to the virus with high rates of fatalities [2], particularly in some residential care homes [3,4] during the first wave of the virus [5]. Hospitalisation rates were also high among those living with long term conditions (LTCs)[2,6], many of which affect the older adult population [7,8]. The UK government imposed social distancing restrictions on March 23^rd^, 2020, where adults over the age of 70 were required to self-isolate and “lockdown” at home for three months to reduce their infection risk.

Drawing on evidence of negative psychological responses during previous epidemics[9], concerns rose among stakeholders at the start of the pandemic that there would be adverse effects of the COVID-19 pandemic on mental health and wellbeing. Whilst under usual circumstances, older adults do tend to experience psychosocial wellbeing that is equal or better than that of younger age groups [10], it was predicted that due to the specific isolation rules for older adults and their heightened risk from the virus, psychosocial consequences such as loneliness would be exacerbated in older age groups [11], leading to negative effects on mental and physical health [12,13]. At a population level mental health during the COVID-19 pandemic was negatively impacted by COVID-19 [14], but older adults on average experienced more stable and less negative outcomes compared with other subgroups [15,16]. It is presently unclear why this was: which underlying factors accounted for the positive and negative experiences reported by older adults during lockdown.

Several theories could help explain the apparent psychological resilience of older adults during the pandemic. Offers of support from social contacts [17], a stable living environment [18], cohabiting with others [19], and financial security[16] may have helped protect this subgroup against adverse effects of social distancing measures by providing a buffer against distress. Additionally, older adults may have drawn on previous life experiences to see a greater sense of coherence in the events of the pandemic. Coherence incorporates comprehensibility (ability to understand and integrate), manageability (ability to navigate and manage) and meaningfulness (sense making) in relation to a new health threat [20]. It has been shown to support better navigation of life stressors [20] and is a strong predictor of health status among older adults [21]. Life wisdom accumulated by older age has also been found to increase the use of problem-focused coping skills and perceived control over events, both of which protect against distress [22].

However, whether factors such as these do indeed explain the better responses amongst older adults remains unexplored. Understanding of these factors that are transferable across groups is essential in developing future interventions and policy for those most at risk of harm due to social distancing measures. Further, whilst the average mental health symptom scores and wellbeing levels of older adults were better than amongst younger age groups, this does not necessarily imply that older adults were therefore unaffected psychologically during the pandemic. Consequently, this study explored in detail the experiences of older adults living in the UK, with two specific research questions: (1) How was the mental health of older adults affected during the pandemic? (2) What might have protected mental health in older adults during this time?

## 2. Methods

### 2.1 Study Design

As part of the COVID-19 Social Study (CSS) that began on March 21^st^ 2020 [23], which is the largest UK study into the psychosocial impact of the pandemic, we undertook a substudy to elicit experiences of older adults. A phenomenological approach was used to interrogate the data, with an idiographic focus on personal accounts of experience. The UCL Ethics Committee reviewed and approved this study (Project ID: 14895/005). Content in the following sections are informed by the COREQ reporting guidelines [24].

### 2.2 Recruitment

Eligibility criteria included: aged 70 years or older, and the ability to speak English sufficiently to understand the study Participant Information Sheet (PIS) and consent form. We recruited participants by listing the substudy in the CSS newsletter (reaching 3,919 subscribers), social media, and through two community organisations who circulated study information within their networks. We used purposive sampling methods to recruit 20 adults aged over 70 with diverse characteristics such as gender, ethnicity, marital status, and living situation. Recruitment ended after 20 one-off interviews, as the lead author AM identified no new themes during the analysis.

### 2.3 Procedure

People interested in participating were asked to contact the research team directly via email. A researcher (AM or AB) responded with further details about the study and an invitation to ask additional questions. All participants then provided written informed consent prior to attending a remote interview by telephone or video call. Participants were offered a £10 shopping voucher as an expression of gratitude. A team of experienced postgraduate-level qualitative female healthcare researchers (AM, AB, LB, AR, SC) conducted one-to-one interviews between May and September 2020. No researcher had prior relations with any research participant. Interview times ranged from 16 to 85 minutes and lasted for 50 minutes on average. A complete interview guide can be found in the Appendix. In brief, interview topics included: normal life before the pandemic, understanding of social distancing guidelines, social life, mental health, and prospection (for question examples, refer Figure 1). Interview guide development was informed by existing theories on social networks and health [25], and sense of coherence theory [26].

**Figure 1.**
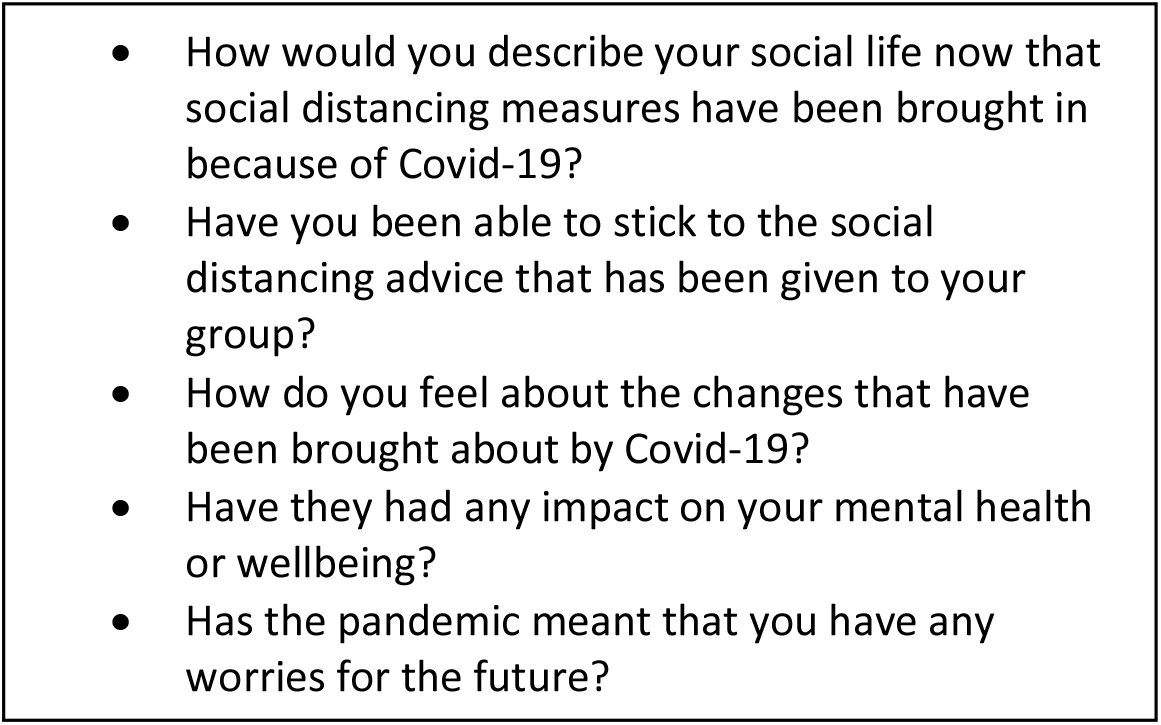
Example of Interview Guide Questions.

**Figure 2.**
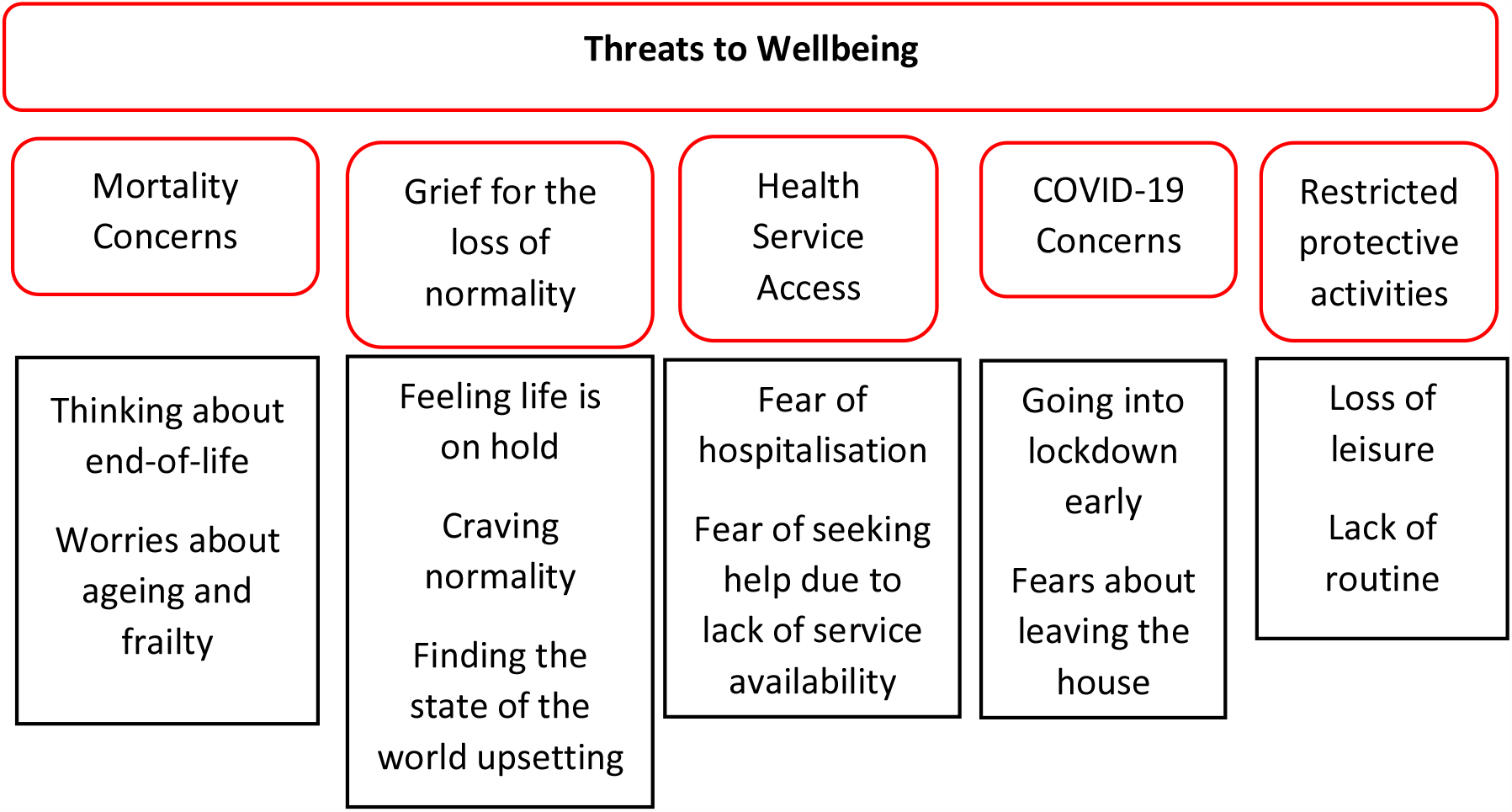
Commonly reported themes about threats to wellbeing during lockdown.

### 2.4 Data Analysis

Researchers audio recorded the interviews with consent from participants, which were then transcribed verbatim by a professional transcription service. All transcripts were double checked for anonymity after transcription before importing into Nvivo version 12 for analysis. For consistency of coding approach, AM and AB double coded 3 transcripts at the start of data analysis and discussed issues of salience raised by participants. The lead researcher (AM) used an inductive and deductive thematic analysis approach, informed by Braun and Clarke [27,28]. An initial coding framework was established based on the supporting theory [25,26] and interview topic guide. This framework was applied to each transcript through line-by-line coding, then the framework was updated with new codes as AM identified new concepts in the transcripts. Themes and subthemes were developed based on participant narratives, and these were presented to the CSS research team on 3 occasions for formative feedback.

## 3. Results

Of those who agreed to take part, 9 participants were women and 11 men, aged 72 to 93 (Table 1). Fourteen reported a physical health condition, including hypertension, diabetes, arthritis, high blood pressure and cancer. Two participants had an anxiety-related mental health condition, and 3 said they had caregiving responsibilities for a spouse or family member.

**Table 1.** Sample characteristics

| Demographic factors | Sample<br>*n* = 20 |
| --- | --- |
| **Age bands** |  |
| 70s | 10 |
| 80s | 9 |
| 90s | 1 |
| **Ethnicity** |  |
| White British | 16 |
| Asian Other | 2 |
| Other (Myanmar) | 1 |
| White Other | 1 |
| **Relationship status** |  |
| Married/Partnership/Cohabiting | 10 |
| Widowed | 6 |
| Separated/ Divorced | 3 |
| Single (never married) | 1 |
| **Employment** |  |
| Retired | 16 |
| Working part time | 2 |
| Self employed | 2 |
| **Living situation** |  |
| With partner/spouse | 9 |
| Alone | 8 |
| With family | 2 |
| With friends | 1 |

### 3.1 Threats to Wellbeing

Some participants described a transient period of uncertainty at the start of lockdown, associated with nervousness and lack of sleep that resolved quickly. Many were concerned about the impact the pandemic was having on their end-of-life experience and the rest of the world. A commonly reported concern among participants was a fear of needing healthcare assistance during the COVID-19 lockdown. Some participants were concerned about catching COVID and additional risks due to their age, and others said that lockdown measures meant they were unable to engage in activities that usually formed part of their self-care routine.

#### 3.1.1 Concerns about End-of-Life, Ageing, and Mortality

More than half of the group spoke about how the pandemic caused them to think about their experience of ageing, end-of-life, and mortality.

> *“It’s just this idea of all of a sudden realising that I’m getting really old. I think that may be the biggest thing, and it’s a combination of getting really old, and the pandemic is probably accentuating it a bit*.*” p10, female, aged 75-79*

Some planned for the possibility of becoming unwell from COVID. For instance, one participant had instructed his family to “stay away” should he become gravely ill, to protect them from the virus. For several participants, a reminder of their own mortality risk came from knowing someone who had passed away from COVID:

> *“We’ve had one friend who was in his sixties… Suddenly went into hospital went on a respirator and sadly he died. He’s the only person we know who has directly been affected by it. It hits you and it makes you realise your own mortality. Especially when they keep saying it affects older people worse, so you do worry*.*” p13, male, aged 75-79*

Several participants described concerns about their vulnerability to COVID due to their age:

> *“It is scary for us at our age. The thought of getting COVID that really frightens me and frightens me for anybody close to me that if they got it. It really terrifies me. So, we have been very, very careful*.*” p11, female, aged 70-74*

#### 3.1.2 Grief and Loss of Normality

A longing for normality was frequently described by participants. Some felt the activities they previously enjoyed, like travelling, going to the theatre, or “hitting a tennis ball,” would never return to the normal they were used to. Others said they felt their life was on hold until the virus was under control or a vaccine was introduced.

> *“The new normal is not going to be at all like the old normal, I don’t think. We won’t really be able to live the kind of life that we lived before until there’s a vaccine, and it looks as though the vaccine is going to be a very long way off*.*” p4, female, aged 70-74*

Some said they felt grief about the impact COVID was having on the world, particularly regarding death, hardships, and suffering of others.

> *“I knew of the wars and the disease and the hunger, but I think COVID has just put a whole blanket round the lot of it and makes it so immense, the state of the world. The horrible state of the world and that is very depressing when you think about it*.*” p19, female, aged 80-84*

#### 3.1.3 Restricted Healthcare Access during Lockdown

Concerns about catching COVID were variable among the group, but many were fearful of being hospitalised for any reason because they believed they were at increased risk of death.

> *“A lot of people are scared stiff of catching [COVID], I’m not. The only thing I’m scared of is being carted off to a hospital. I want to die peacefully at home, and I would happily do that any night*.*” p8, male, aged 90-94*

Some worried about the lack of available health services during lockdown, particularly should anything “go wrong” with their health independently from COVID.

> *“A friend of mine has just been diagnosed with breast cancer. She’s had to wait about nine weeks for her op… so you worry about if something like that happened to me, would I get the medical attention I need?” p12, female, aged 75-79*

#### 3.1.4 COVID-19 Concerns

The potential health threat of COVID meant some participants were scared to leave the house:

> *“I do feel that perhaps I should be going out more and that sort of thing, but myself and many, many, almost all my friends say that they are very scared to go out*.*” p2, female, 70-74*

Several said they did not think a COVID vaccine would be available to help life “go back to where we were before.”

> *“Whatever happens, even if a vaccine comes, we will never return to shaking strangers’ hands*.*” p16, male, aged 75-79*

#### 3.1.5 Unable to engage with activities that protect wellbeing

Due to social distancing and travel restrictions, some participants were unable to engage in activities such as weekly religious ceremonies, theatre groups, and sports. Although some activities were able to be online, this was not always possible.

> *“Since COVID, [community activities have] all closed down. Well yes, the book club totally because we can’t discuss books over the phone and also people are of an age where you can’t do social media, whatever you call it*.*” p19, female, aged 80-84*

Several participants commented on the consequences of a disrupted routine on their wellbeing during lockdown.

> *“That was the first thing that hit me, boredom. I had no idea what the hell am I going to do next, because I was used to a routine and suddenly the routine was completely disrupted…Now suddenly I had nothing to do and I was really lost. I was walking round the house like a bloody zombie trying to find something to do*.*” p15, male, aged 80-84*

### 3.2 Protective Activities and Behaviours

Despite voicing some threats to wellbeing, participants were mostly positive in reflecting on their lockdown experience overall. This was attributed to a slowed pace of life, maintaining a routine, using coping skills and resources, and accessing social support (Figure 3).

**Figure 3.**
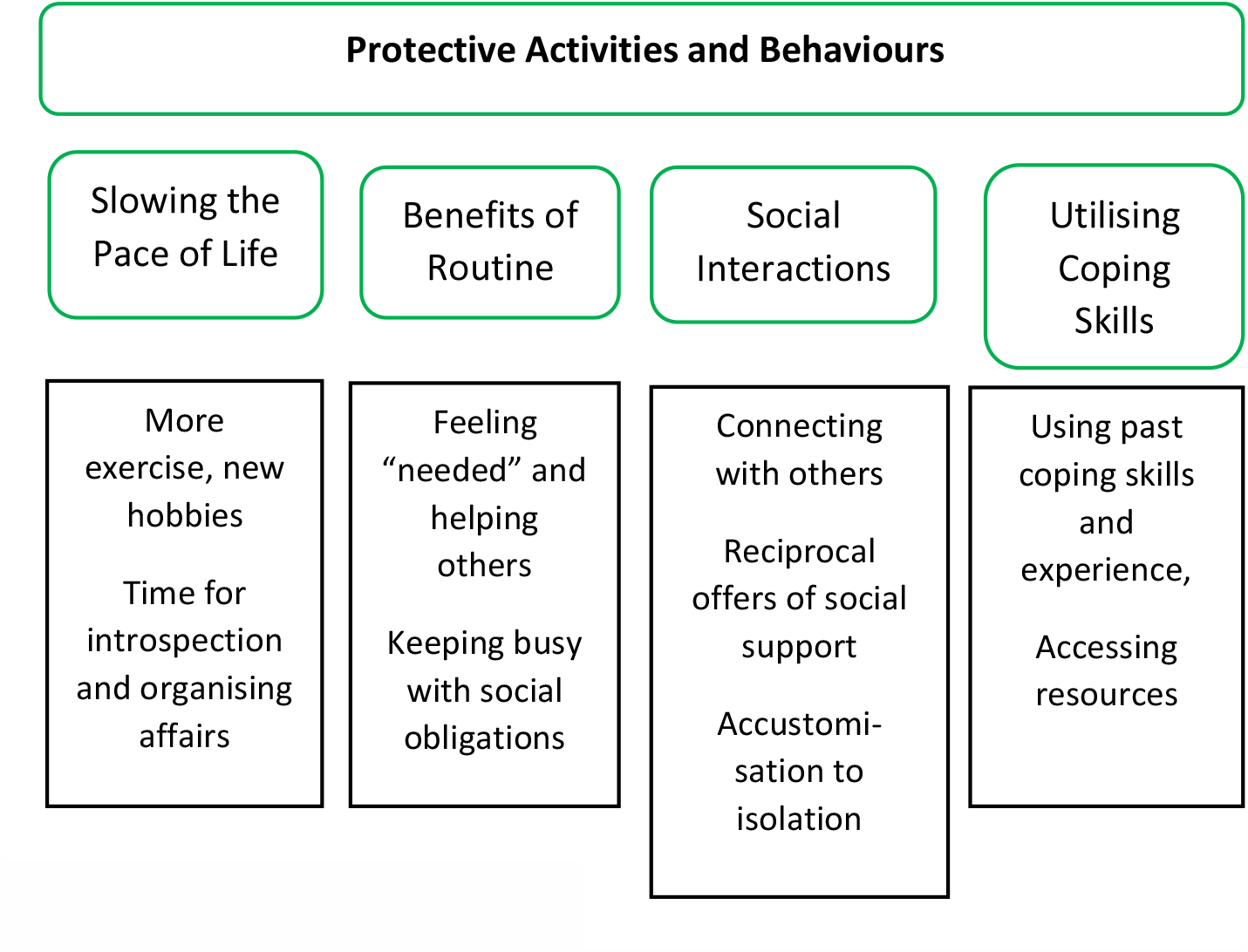
Commonly reported themes about protective aspects of lockdown experience.

#### 3.2.1 Slowing the Pace of Life

The most commonly reported experience during lockdown was feeling like the pace of life had slowed with more time to reflect. Although some participants had described a loss of leisure during lockdown, many had actually found time for new hobbies, reading, crafts, gardening, and learning a new language.

> *“Sometimes I wake up in the morning, and I think, oh, it’s another day in lockdown, but I think… there are some little positive benefits…Before lockdown, we were all rushing around doing lots of things, and now we’ve had to slow down. And actually, slowing down has been quite nice. And we’re living in the kind of retirement now that, maybe, our grandparents might have lived, when you just cultivate your garden and do your knitting and crochet*… *But just generally living a slower pace of life*.*” p4, female, aged 70-74*

Others felt that being required to stay at home presented an opportunity to focus more on their health by going for regular walks and taking up new forms of physical activity. For some, this was the first time in decades they had been so physically active.

> *“I’ve now started to ride a proper bike as well. I live in a Close, so we don’t get any through traffic and I can cycle around that Close and I do a few laps. But I haven’t ridden a bike for 60 years*.*” p1, male, aged 80-84*

Half of the group said the slowed pace of life gave more time for introspection, with many reflecting on their life differently and in a more meaningful or positive way than they had before: *“I’m not rushing around so much anymore, it’s given me the time and the opportunity to notice small things*.*” p4, female, aged 70-74*. Many used this process of reflection to think about the changes they would make to their lives as a result of their pandemic experience.

> *“Having grown quite a lot, I feel quite positive about that. I also think I’m going to try and, maybe, achieve more things when I come out of this [lockdown]. I think when you retire, and as you get older, you become very comfortable in your life. I think, perhaps, I was a bit too comfortable. I need to get out and be more proactive*.*” p6, female, aged 70-74*

#### 3.2.2 Benefits of Routine and Social Responsibilities

Nearly half of participants said that maintaining a routine and sense of purpose was important for their wellbeing during the COVID-19 lockdown: *“You have to have a purpose you see. I think mental resilience is all about having a sense of purpose*.*” (p15, male, aged 80-84)*.

Some said they experienced a meaningful “psychological benefit” from social responsibilities, such as cooking a meal for family, phoning friends to check in, or caring for a pet:

> *“The important thing is to have the necessity to do things. Whether it is to get in touch with people, to write a piece of something… Obligations are a good source of maintaining ones feeling of self-worth, if you like. So I think it’s very important to make sure that whatever it is, even though you may feel oh what a nuisance I’ve got to do that, the very fact of having to do it is a great psychological benefit*.*” p3, female, aged 70-74*

#### 3.2.3 Social Interactions and Support

The nature of socialising had changed since the start of lockdown for many but not all participants. Several said they were socialising to try to carry on *“life as normal”*, particularly keeping in regular contact with family. For some, this resulted in strengthened relationship bonds and connectedness:

> *“I think it has made me and my husband stronger really. We’ve never spent as much time together actually… I think we’ve coped with the shopping and organising that. And we’ve been baking together, we’ve never done things like that. And we took it in turns to cook and tidy up after. We have done really well together. I’m really proud of us*.*” p11, female, aged 70-74*

For some who lived alone, they spoke of being accustomed to isolation long before the pandemic arrived: *“I’m a fairly sort of isolated person anyway*.*”* P1. Several said they were accustomed to being alone due to widowhood or retirement, and therefore lockdown did not prompt a dramatic change in their daily living or social life:

> *“I’ve been retired for a nice long time… So, in many ways the lockdown, on one side it hasn’t impacted a great deal, because I was used to being at home and certainly over the past two years to being home alone*.*” p20, male, aged 80-84*

#### 3.2.4 Utilising Skills, Experience and Resources to Cope

Participants who had used mental health services in the past spoke of utilising the skills they had learned to cope with the COVID-19 crisis, including mindfulness and meditation.

> *“I had a wonderful counsellor who I saw about once a year, and she would set me on the right path. And eventually, after many years of trying, I found a mindfulness and meditation book, about the middle of last year… so I feel that that has been a great help to me. Usually I try in the morning and certainly in the evening, before I go to bed, I do some meditation*.*” p2, female, 70-74*

Others described experiences of hardship in the past that they used as a comparison with COVID times, such as living through war, displacement, and illness:

> *“I was diagnosed with what they call non-invasive bladder cancer… Having gone through the concern of something like that, perhaps Covid, you know, you put it into perspective*.*” P13, male, aged 75-79*

Participants frequently mentioned their access to resources that had been arranged during lockdown, with support offered by family, friends, and neighbours.

> *“I’ve had online shopping every week since lockdown and I haven’t been to any shop. Prescriptions were delivered and anything I wanted, my daughter would fetch*.*” p18, female, aged 80-84*

## 4. Discussion

In this study, we sought the views of older adults about factors that threatened or protected their mental health during the COVID-19 pandemic. Although in previous research older adults statistically experienced better mental health than other age groups during the pandemic [30], our study identified a number of threats to wellbeing in this age group, including fears relating to the virus, the future, and their own mortality. However, older adults also described a range of activities and behaviours that helped to protect their mental health and could be used to explain their better levels of mental health and wellbeing relative to other age-groups. Congruent with international research [17], many participants began to self-isolate earlier than guidance required and perhaps consequently, arrangements were in place for access to essentials from the outset of lockdown, resulting in greater sense of coherence of COVID-19 as a potential health threat. For the most part, participants enjoyed less social pressure and more time for hobbies. Similarities in experience were drawn between a slower pace of life germane to retirement and day-to-day realities of the COVID-19 lockdown.

### Factors that threatened mental health during COVID-19

Given early evidence publicised on mortality risk for older adults[31], it is unsurprising participants frequently discussed concerns about their end-of-life. Some were worried about the physical and cognitive impact of lockdown on their experience of ageing. Studies have shown an association between social isolation and reduced physical performance [32], which caused concern among some participants in our study and resulted in many taking extra steps to preserve activity levels. Whilst this may have provided positive health benefits in the short term, of most concern is the fear many participants described in leaving the house to access routine or preventative health care, which may have longer-term implications for public health services if this age group experiences more physical health challenges in the coming months. Aligned with international research [33], participants in our study also worried about the impact of COVID-19 on the world and spoke of the impact this had for their wellbeing on a daily basis. Feelings of grief and loss were frequent in this group and is likely to be felt across many societies in response to the pandemic.

### Factors that protected mental health during COVID-19

Quantitative data collected during the first UK lockdown suggests that those with restricted finances and access to basic needs experienced higher levels of adversity during the first wave of the pandemic [34]. Many participants in our study reported having access to basic supplies, high levels of perceived social support, and secure living arrangements, which helped to create a buffer for them against stress and uncertainty. National averages showed infrequent experiences of loneliness among older adults [35], which may be explained by our finding that participants engaged frequently in online methods of interaction, spent time with pets, and/or had regular “check-ins” with friends and family to mitigate against loneliness. As such, the heightened concern about loneliness in this age group early on in the pandemic may have led older adults and those around them to proactively take steps that helped prevent these experiences in many individuals. Indeed, many participants reported enhanced feelings of connectedness with social contacts throughout the lockdown, which can prevent isolation and protect against emotional distress [12]. However, a small number of participants did not feel connected, particularly those who had been separated from their family because of the pandemic, highlighting that when such support was not available, it was related to less positive wellbeing experiences.

### Implications

This study highlights a number of important implications. First, the threats to wellbeing amongst older adults need addressing as they have implications for the immediate future and for future pandemics. In trying to remove barriers to healthcare access, supporting older adults in engaging with telecare may be a helpful alternative for some health concerns. However, in our work involving people with mental health conditions, we found service users felt this was often an unhelpful substitute [36]. Future research must address indirect health consequences of the pandemic resultant of delayed or diminished access to healthcare during the lockdown. Second, as discussed elsewhere [37], interventions to mitigate the impact of prolonged isolation on experiences of grief are warranted. Grief can prompt search for meaning and seeking out others with similar experiences. Clinicians play a role in supporting people in processing their grief associated with COVID, but spaces online and within groups may also facilitate healing from loss experienced during the pandemic [37]. Schemes such as social prescribing could be deployed to support older adults psychosocially, and may provide additional support in the aftermath of COVID-19 [38]. Finally, it is evident that forward planning by families and communities to address initial concerns about older adults during the pandemic played an important role in supporting their coping and buffering against loneliness, isolation, and poor mental health. For future pandemics, such response is again encouraged. In particular, interventions that bolster feelings of certainty and connectedness may serve as helpful targets for those experiencing pandemic-related distress.

### 4.1 Strengths and limitations

A strength of this research is that data were collected from participants via purposive recruitment throughout the first UK lockdown and as restrictions began to ease before the second wave. However, findings must be interpreted cautiously. Our participants were generally healthy, with well-established social networks, living in the community, and predominantly without solo caregiving responsibilities. Therefore, their experiences are not likely to be representative of those living with serious health concerns, who may be more likely to have experienced distress during the pandemic [39]. We conducted interviews via video call or telephone, which meant being able to capture experiences safely amid restrictions, but also means that those without access to the internet or telephone would not have had equitable access to participate and may have faced additional challenges. We also did not collect data on, or sample based on previous COVID-19 infection and to our knowledge, none had experienced a confirmed diagnosis. So future studies are needed to ascertain how older adults who experienced COVID-19 were affected psychologically [35].

### 4.2 Conclusions

Contrary to early concerns at the start of the pandemic, the mental health of older adults fared well compared with other age groups, and this study adds to the literature on this topic by providing evidence as to why these results may have been found. Overall, many participants described their experience of lockdown as a time for reduced social pressures and personal growth. However, this group were not without challenge, particularly among those who were concerned about staying well, advancing frailty, or hospitalisation risk. This research therefore highlights the importance of nuance when considering the relative better experiences of older adults. It also provides valuable insight into factors that protected wellbeing of older adults during the COVID-19 pandemic, which may be utilised by policy makers to support at-risk groups who have experienced psychological hardship during the crisis, including timely access to essential supplies, communicating offers of help to improve perceived social support, and providing structure and routine in times of uncertainty.

## Data Availability

No research data are publicly available as this may contain information that could affect research participant anonymity.

## Ethical approval

The study was reviewed and approved by the UCL Ethics Committee (Project ID 14895/005).

## Acknowledgements

The researchers are grateful for the support of AgeUK, the Alzheimer’s Society and Healthwise Wales during recruitment. Many thanks to Anna Roberts, Louise Baxter and Sara Conway for their help with conducting interviews. Thank you to the COVID-19 Social Study Team (Tom May, Katey Warren, Joanna Dawes, and Henry Aughterson) who provided feedback on the themes and subthemes.

## Appendix Interview Guide for Adults

